# Quantifying hospital flows and occupancy due to COVID-19 outbreak in France. Was French lockdown effective?

**DOI:** 10.1101/2020.06.08.20125765

**Authors:** David Ternant

**Affiliations:** EA 7501, Université de Tours, Tours, France

## Abstract

**Context:** The spread of SARS-CoV-2 led to a rapid and deadly pandemic which reached almost all countries in the world in a few months. In most countries, rigorous measures of mitigation, including national or subnational lockdowns were established. The present work aimed at quantifying the effect of national lockdown in France on hospital occupancy.

**Methods:** A compartmental model describing patient hospital flows was developed, where the effect of lockdown was quantified on the hospitalization rate. Model parameters were estimated using nonlinear mixed-effects (NLME) modelling.

**Results:** French lockdown led to a hospitalization rate decreased by thrice from 12 days after its beginning. However, lockdown may not to have decreased either hospital occupancy or deaths in hospital, which would have been both decreased by 30% and 85% in average if lockdown was started 20 and 30 days before this date, respectively.

**Conclusion:** The present work showed an intrinsic effect of lockdown to decrease hospital burden, but its efficacy on hospital burden may have been increased if established sooner.

## 1 Introduction

After an outbreak in December 2010 in Wuhan (Hubei, China), Coronavirus Disease 2019 (COVID-19) reached the rest of the world in less than two months. The World Health Organization (WHO) declared on February 26 of 2020 that all continents were touched by COVID-19 and declared pandemic situation on March 11. On May 22^nd^, more than 5 million of cases and 330000 of deaths were recorded: most of deaths were recorded in USA (96363), United Kindgom (36042), Italy (32486), France (28215), Spain (27940) and Brazil (20082) [1].

The report 9 of Ferguson et al. [2] suggested that COVID-19 may trigger not only dramatic number of deaths, but also huge hospital overload and saturation of intensive care units (ICU). Therefore, 87 countries have imposed national or subnational lockdowns [3]. In France, the first COVID-19 cases were confirmed at the end of January, and the first death from COVID-19 was recorded on February 28; a national lockdown, including overseas territories and departments, was begun on March 17^th^ and ended on May 11^th^ in France [4].

Previous works that used compartmental deterministic mathematical models suggested and/or confirmed substantial benefit of lockdown [5–9]. The validity of compartmental deterministic models by themselves is usually agreed. However, since they are most often used as infection and/or death count forecasts very early in epidemic histories, their (numerous) parameters have to be fixed based on prior knowledge, which might lead to over- or under-predictions.

Compartmental models have been used in several fields, including medical research and pharmacology, where compartmental models are usually estimated from data to quantify biologic and/or pharmacologic phenomena, while no (or a few) parameter values are fixed from assumptions. In this field, non-linear mixed-effects (NLME) modelling is used to estimate model parameters with methods based on likelihood maximization or Markov-chain Monte-Carlo (MCMC) methods [11–12].

If most of previous works used sound epidemic and/or mechanistic models to describe COVID-19 outbreak, spread and hospital occupancy, none of them attempted to estimate models parameters to quantify actual patient flows in hospital services and ICU or the influence of lockdown on hospital occupancy. Therefore, the present study proposes to develop a compartmental model describing hospital flows between hospital services, ICU, deaths and return home, to estimate transfer flow rate constants, and to quantify the effect of lockdown in France using NLME modelling.

## 2 Methods

### 2.1 Data extraction

The present work was made on French data which were collected from official databases [4]. Data that were considered were hospitalizations in (i) non-ICU units and (ii) in ICU units, (iii) deaths in the hospital, and (iv) returns to home. These data were collected daily from March 1^st^ towards May 22^nd^ of 2020. They were assessed by regions (or sub-regions) selected to keep comparable number of patients within each unit. Indeed, regions were divided in smaller areas if effectives were high, as in Ile-de-France, Grand-Est, Hauts-de-France, Auvergne-Rhône-Alpes and Provence-Alpes-Côtes d’Azur. Therefore, the following 26 regions or sub-regions were assessed : Paris, Hauts-de-Seine, Val-de-Marne, Seine-saint-Denis, Essonne, Yvelines, Val d’Oise, Seine-et-Marne, Haut-Rhin, Bas-Rhin, Moselle, Grand-Est (other), Nord, Hauts-De-France (other), Rhône, Auvergne-Rhône-Alpes (other), Bouches-du-Rhône, Provence-Alpes-Côtes d’Azur (other), Bourgogne-Franche-Comté, Centre-Val-de-Loire, Occitanie, Nouvelle-Aquitaine, Pays-de-la-Loire, Normandie, Bretagne, Corse. Overseas departments and territories were not assessed because of low effectives. No focus was made on COVID-19 cases, because no systematic testing policy was implemented in France up to this date. Since COVID-19 cases were not accounted, the model was made under the assumption of a constant ratio between cases leading to hospitalization, and total cases.

### 2.2 Hospital flow model

The hospital flow model is described using a system of 9 ordinary differential equations. (figure 1). This model was obtained after having tested numerous possibilities. First, patient hospitalization was described using a Kermack-McKendrick input function:

**Figure 1.**
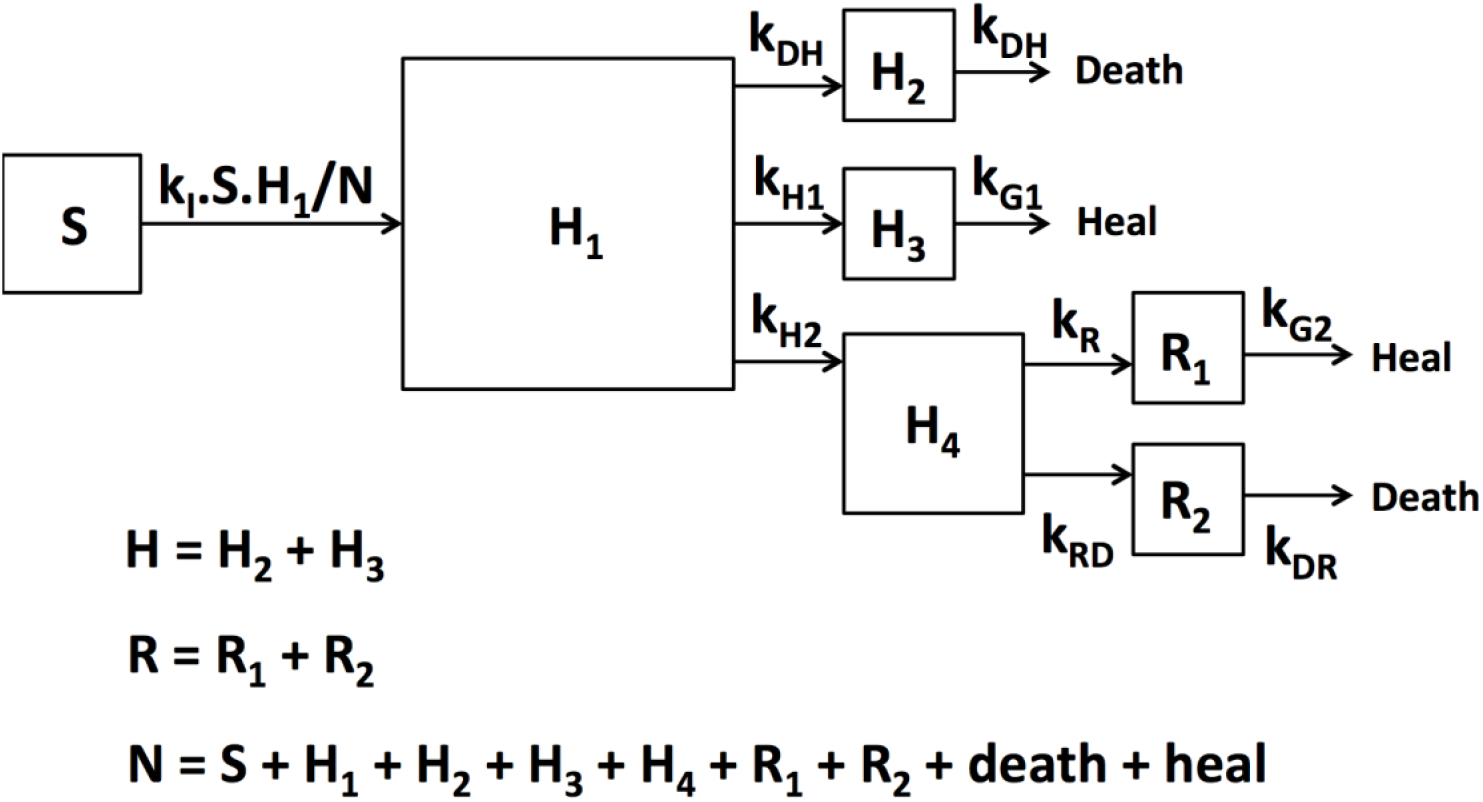
Compartmental model describing patient flows between susceptible (S), non-ICU services (H), in ICU (R), deaths in hospital (D) and return to home (G). Abbreviations are found in the text below.

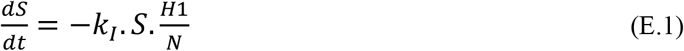

Where S is the number of subjects susceptible to be hospitalized, k_I_ is hospitalization rate constant, H_1_ is the number of hospitalized subjects and N is total number of subjects. Hospitalized patients are assumed to be directed to heal, to die or towards ICU:

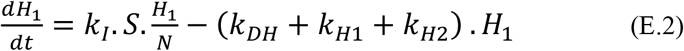

Where k_DH_, k_H1_ and k_H2_ are direction rate constants towards hospital death, hospital heal and ICU, respectively. Because of a delay between hospital admission and death, decease in hospital is described using the following equation:

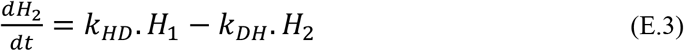

Where H_2_ is the number of hospitalized patients in non-ICU services bound to die, kHD and kDH are input and output rates constants, respectively. Because early attempts showed that patients bound to live and to die present different kinetics, they were described using separate equations. Therefore, the kinetics of patients that heal from hospitalization is described as follows:

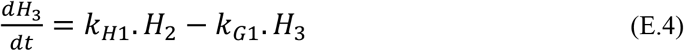

Where H_3_ is the number of hospitalized patients (not in ICU) bound to heal. Early attempts showed a delay between admission in hospital and in ICU. In addition, as for hospital patients, patients bound to heal or die in ICU present different kinetics and were therefore described in separate equations. Thus, the following compartment described the transfer of patients toward ICU as follows:

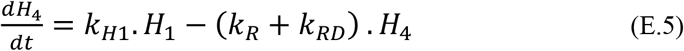

Where H_4_ is the number of patients in tansit compartment which describes a delay of patient direction towards ICU, and k_R_ and k_RD_ are direction rate constants toward heal or death paths in ICU, respectively. The kinetics of atients bound to heal for ICU is described as follows:

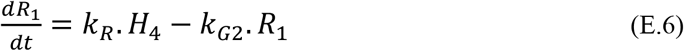

Where R_1_ is the number of patients bound to heal in ICU and k_G2_ is ICU heal rate constant. The kinetics of patients bound to die in ICU is described as follows:

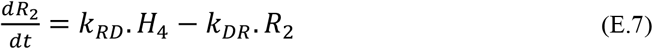

Where R_2_ is the number of patients bound to die in ICU and k_DR_ is ICU death rate constant. Dead (D) and healed (G) patients were described using the following equations:

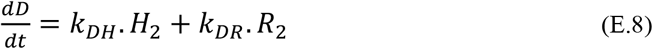

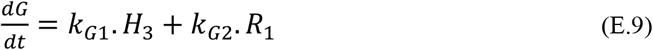

Initial time (t=0) was fixed on the December 31^st^ of 2019, to set January 1^st^ of 2020 as day 1. The system was initialized by estimating the time of the first patient to be hospitalized, i.e. t_first_ such as H_1_(t=t_first_) = 1. All other equations were assumed to be equal to 0 at t = t_first_. The number of hospitalized patients outside ICU (H), in ICU (R) were respectively assumed to be H = H_2_ + H_3_ and R = R_1_ + R_2_. The influence of French lockdown was assumed to decrease the hospitalization rate constant as follows:

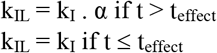

Where α is a coefficient between 0 and 1 and t_effect_ is the time from which lockdown effect was detected.

### 2.3 Parameter estimation

#### The following 14 parameters were estimated

t_first_, N, k_I_, k_HD_, k_DH_, k_H1_, k_H2_, k_G1_, k_R_, k_RD_, k_DR_, k_G2_, α and t_effect_. Parameter estimation was made using non-linear mixed-effects (NLME) modelling. This approach has been used since the early 70’s in the field of quantitative pharmacology [10–11]. The main goals of NLME modelling were to determine the distribution of the values of the parameters of interest in a group of individuals, i.e. regions (and sub-regions) of France; data from all regions or sub-regions were computed simultaneously to estimate (i) the “mean” (referred as “typical”) value of each parameter, (ii) the inter-region variability (referred as “interindividual variance”) and (iii) to estimate parameter values for each region using Bayesian methods.

#### Software

Hospital data were analysed using MONOLIX-Suite 2019 software. (Lixoft®, Antony, France), which combines the stochastic expectation-maximization (SAEM) algorithm and a Markov Chain Monte-Carlo procedure for likelihood maximization. To ensure the best possible convergence, a large number of iterations (15000 for K1 and 2000 for K2) were used. K1 and K2 refer to the SAEM procedure of MONOLIX, called “iterative kernels”. During K1, the sequence of step sizes is constant, which allows the exploration of the parameter space. During K2, the step sizes decrease to ensure convergence. Three Markov chains were used. Parameter values of regions, sub-regions and whole France were obtained by computing conditional distribution.

#### Structural model design

Model with and without lockdown effect on k_I_ were compared using Akaike’s information criterion (AIC), defined as: AIC = OFV + 2.p, where OFV is the value of the objective function and p is the number of model parameters to estimate. The use of AIC is based on the parsimony principle aiming at a satisfactory fitting of the data with a small number of parameters. The model with the lowest AIC was selected.

#### Statistical (inter-region) model

The inter-region variability of parameters was described using an exponential model: θ_i_ = θ_TV_. exp(η_i_), where θ_i_ is the estimated individual parameter, θ_TV_ is the typical value of the parameter and η_i_ is the random effect for the i^th^ patient. The values of η_i_ were assumed to be normally distributed with mean 0 and variance ω^2^. The variability of parameter α, assumed between 0 and 1, was described using a probit model.

#### Error model

Additive, proportional and mixed additive-proportional models were tested. For example, the combined additive-proportional model was implemented as follows: Y_O,ij_ = Y_P,ij_.(1+ ε_prop,i_j) + ε_add,ij_ where Y_O,ij_ and Y_P,ij_ are observed and predicted j^th^ marker measurements for the i^th^ patient, respectively, and ε_prop,ij_ and ε_add,ij_ are proportional and additive errors, which are assumed to follow a Gaussian distribution with mean 0 and variances s_prop_^2^ and s _add_^2^, respectively.

#### Model evaluation

Models were evaluated graphically using goodness-of-fit diagnostic plots: observed vs. population (PRED) and individual (IPRED) fitted concentrations; population (WRES) and individual (IWRES) weighted residuals vs. PRED and IPRED, respectively.

### 2.4 Simulations

Typical estimates of model parameters (table 1) were used to simulate the effect of lockdown on H, R, D and G data in time. The value of N_0_ was converted as its value per 100,000 capita (N_0*_=170). The following *scenarii* were simulated: no lockdown, actual lockdown (on the March 17^th^ of 2020), and lockdown started 10, 20 and 30 days before this date.

**Table 1.** Parameter estimates

| Parameter | Structural model ($\theta$) | | Statistical model ($\omega$) | | Error model ($\sigma$) | | |
| --- | --- | --- | --- | --- | --- | --- | --- |
|  | Estimate | R.S.E.(%) | Estimate | R.S.E.(%) | Parameter | Estimate | R.S.E.(%) |
| $t_{\text{first}}$ | 48 | 2.8 | 0.15 | 13.3 | $\sigma_{\text{add,H}}$ | 14.4 | 4.8 |
| $N_0$ | 5660 | 9.0 | 0.43 | 15.2 | $\sigma_{\text{prop,H}}$ | 0.0412 | 2.2 |
| $k_i$ | 0.32 | 10.1 | 0.13 | 14.7 | $\sigma_{\text{add,R}}$ | 8.7 | 2.9 |
| $k_{\text{HD}}$ | 0.0098 | 7.9 | 0.39 | 15.1 | $\sigma_{\text{prop,R}}$ | 0.048 | 3.3 |
| $k_{\text{DH}}$ | 0.00062 | 47.4 | 2.57 | 16.5 | $\sigma_{\text{add,D}}$ | 9.2 | 2.1 |
| $k_{\text{H1}}$ | 0.040 | 5.9 | 0.27 | 16.0 | $\sigma_{\text{prop,D}}$ | 0.0092 | 4.7 |
| $k_{\text{H2}}$ | 0.026 | 7.5 | 0.38 | 14.2 | $\sigma_{\text{add,G}}$ | 38.2 | 2.4 |
| $k_{\text{G1}}$ | 0.071 | 5.9 | 0.33 | 12.9 | $\sigma_{\text{prop,G}}$ | 0.018 | 3.7 |
| $k_{\text{R}}$ | 0.48 | 12.7 | 0.66 | 14.4 | | | |
| $k_{\text{RD}}$ | 0.47 | 15.8 | 0.82 | 14.3 | | | |
| $k_{\text{G2}}$ | 0.086 | 10.5 | 0.59 | 12.8 | | | |
| $k_{\text{DR}}$ | 0.19 | 9.6 | 0.53 | 13.0 | | | |
| **alpha** | 0.29 | 17.5 | 0.67 | 16.8 |  |  |  |
| $t_{\text{effect}}$ | 88 | 1.2 | 1.1 | 15.1 | | | |

**Legends.** $\theta$ , structural parameters; $\omega$ , inter-region standard deviation of structural parameters; $\sigma$ error parameters; R.S.E: relative standard error = standard deviation of estimates / estimate x 100. Other abbreviations are found in the text.

## 3 Results

Data of patient hospitalization in non-ICU services (H, figure 2A), in ICU (R, figure 2B), deaths in hospital (D, figure 2C) and return to home (G, figure 2D) were satisfactorily described by the hospital flow model. Best error models were mixed additive-proportional for all measurements. All model parameters were estimated with good accuracy (table 1). No estimation collinearity was observed. The plots of predicted vs. observed measurements showed good model fitting (figure 3). Individual weighted residuals showed no obvious bias or model misspecification (figure 4). Accounting for lockdown model led to a real improve in data description, with a decrease in AIC of 1057.02 (AIC = 81072.13 and 80015.11 without and with lockdown effect, respectively).

**Figure 2A.**
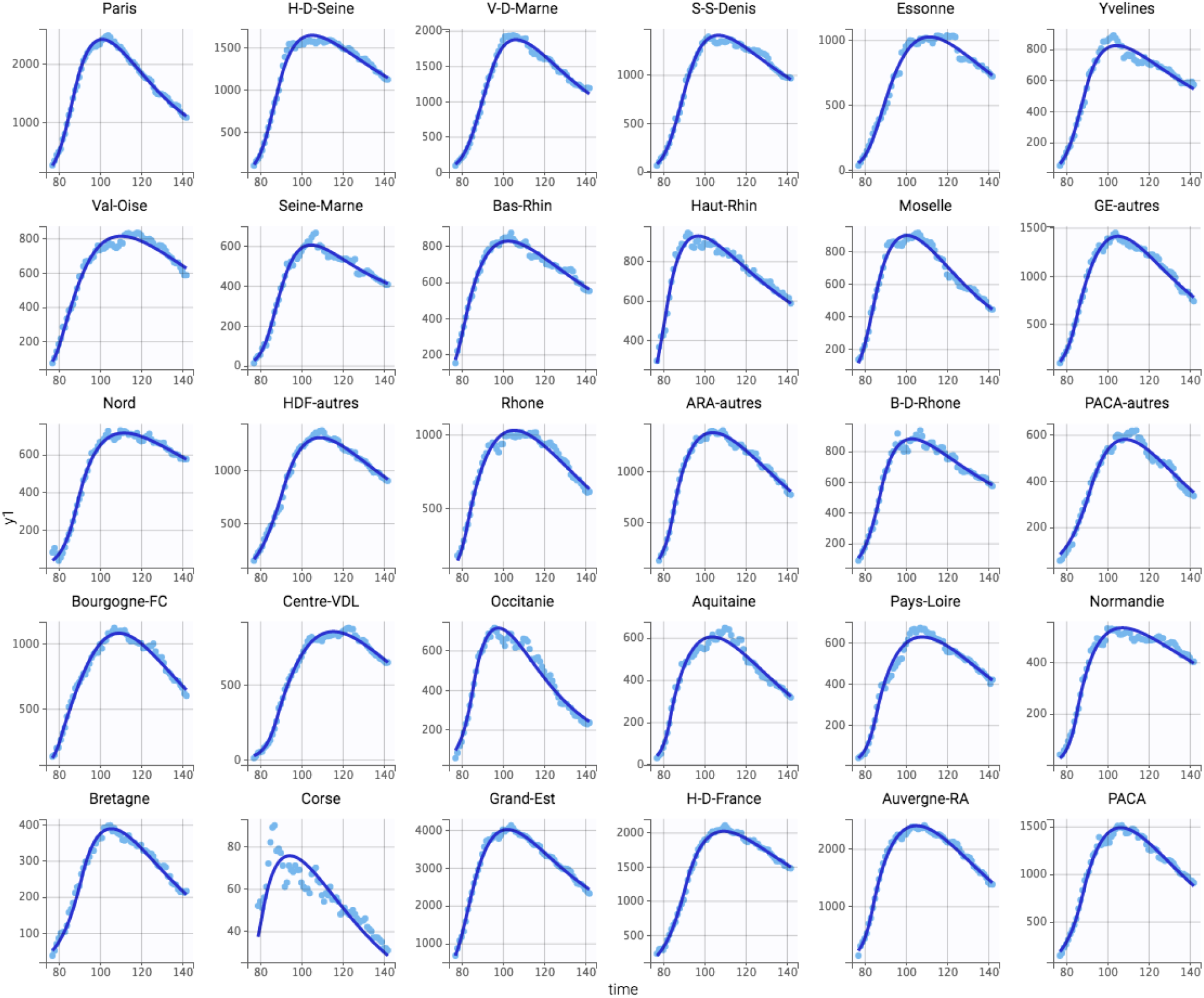
Observed (clear blue dots) and model-fitted (deep blue line) hospitalizations in non-ICU services in different (sub)-regions

**Figure 2B.**
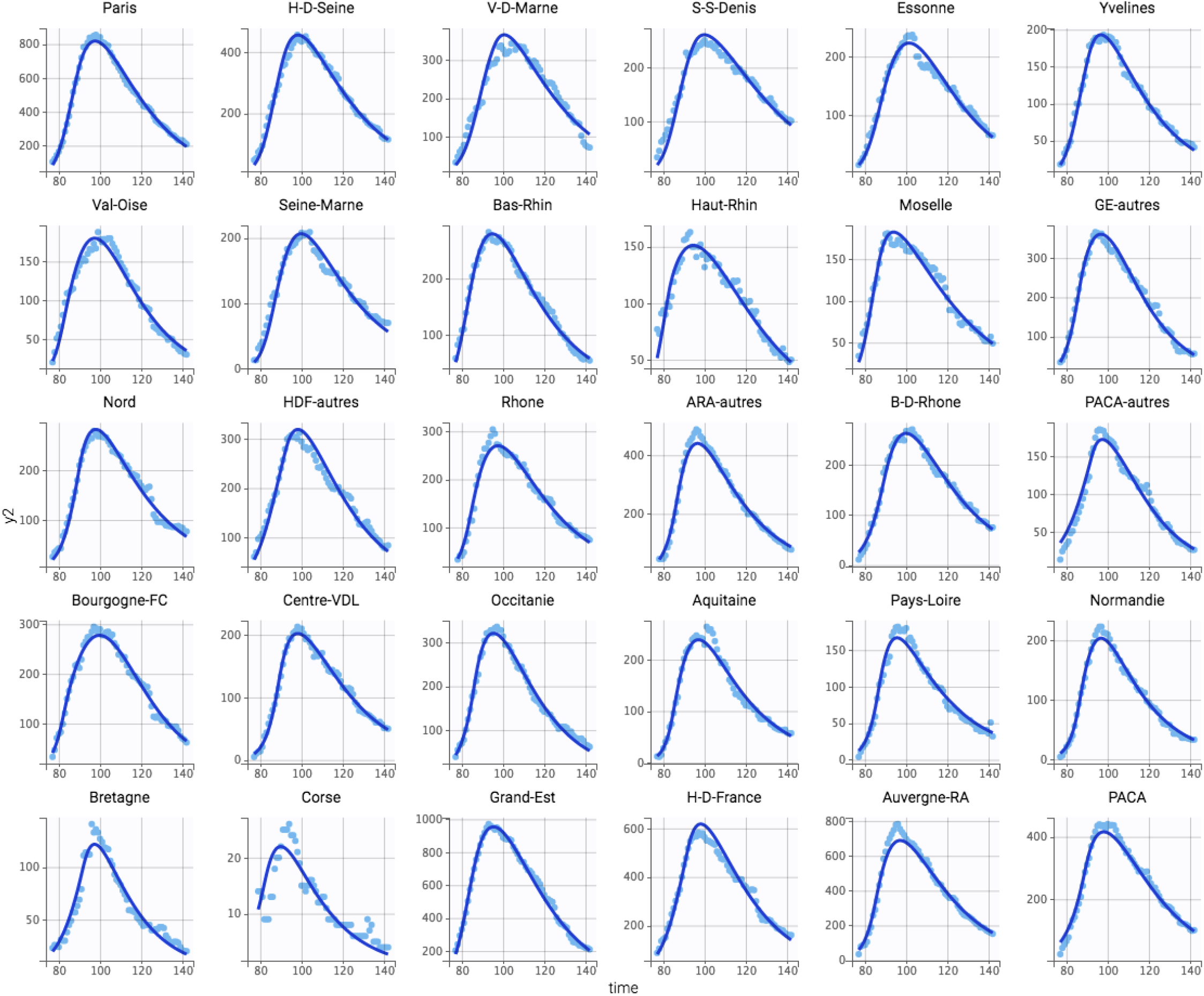
Observed (clear blue dots) and model-fitted (deep blue line) hospitalizations in ICU in different (sub)-regions

**Figure 2C.**
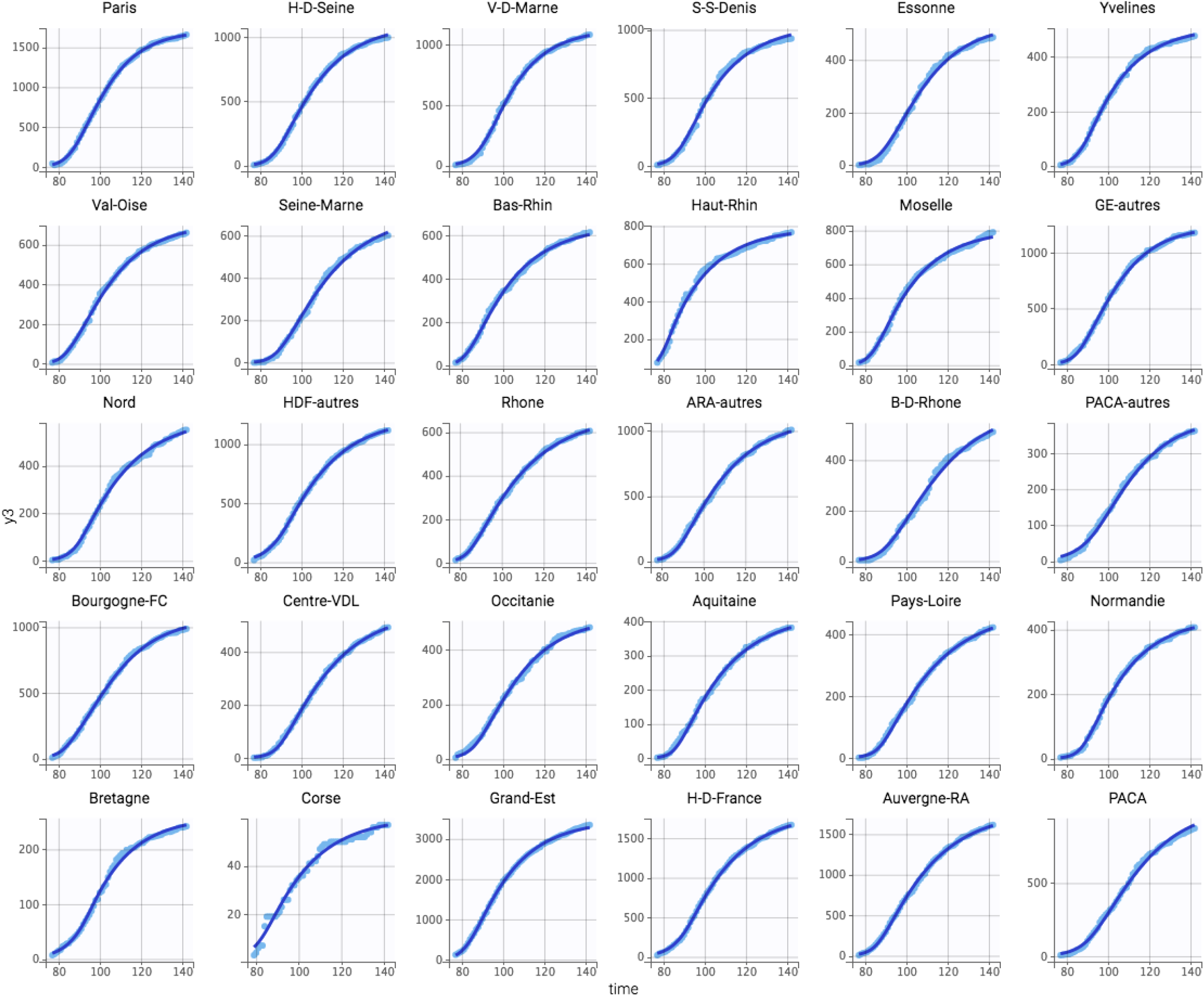
Observed (clear blue dots) and model-fitted (deep blue line) deaths in hospital in different (sub)-regions

**Figure 2D.**
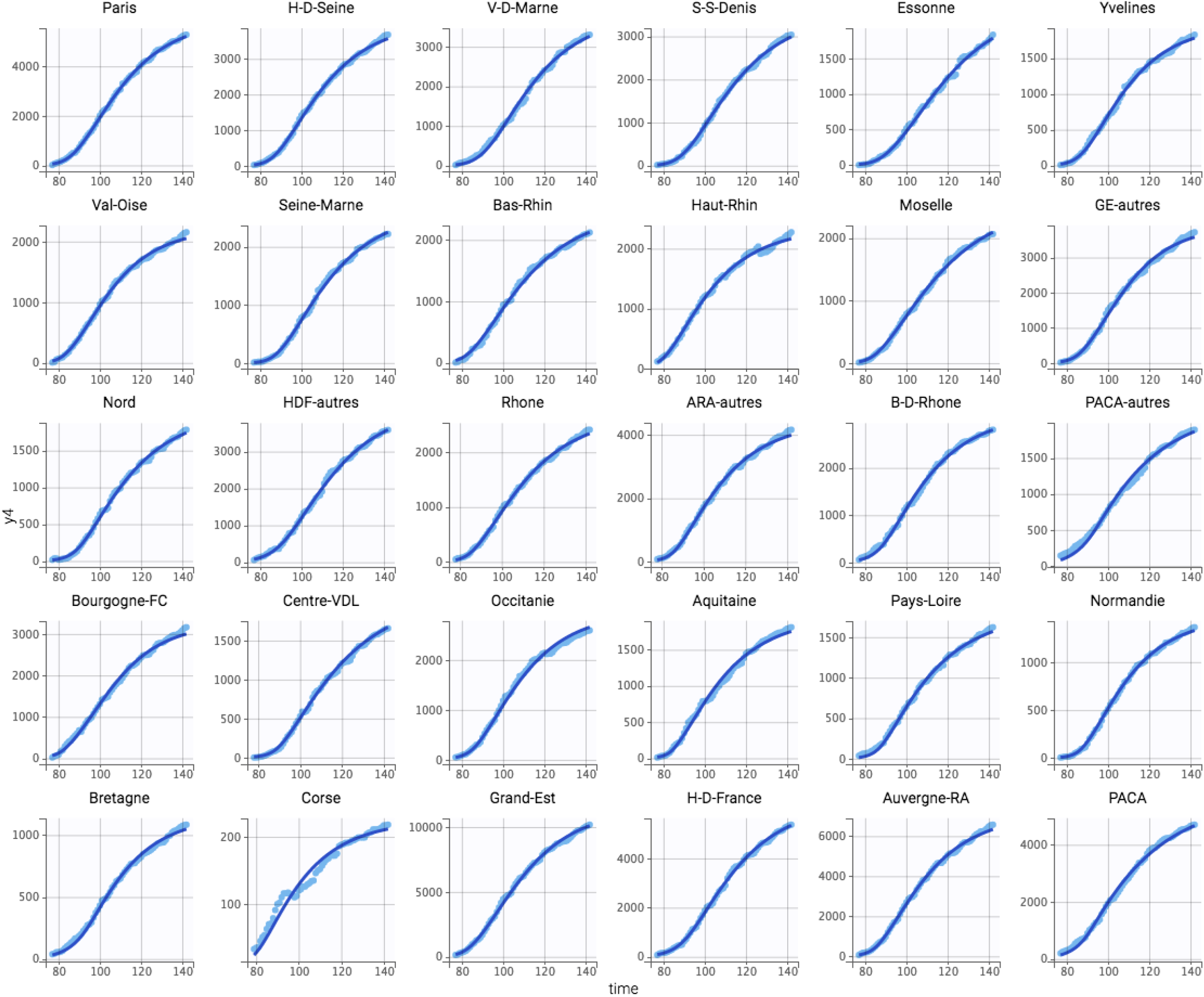
Observed (clear blue dots) and model-fitted (deep blue line) returns to home in different (sub)-regions.

**Figure 3.**
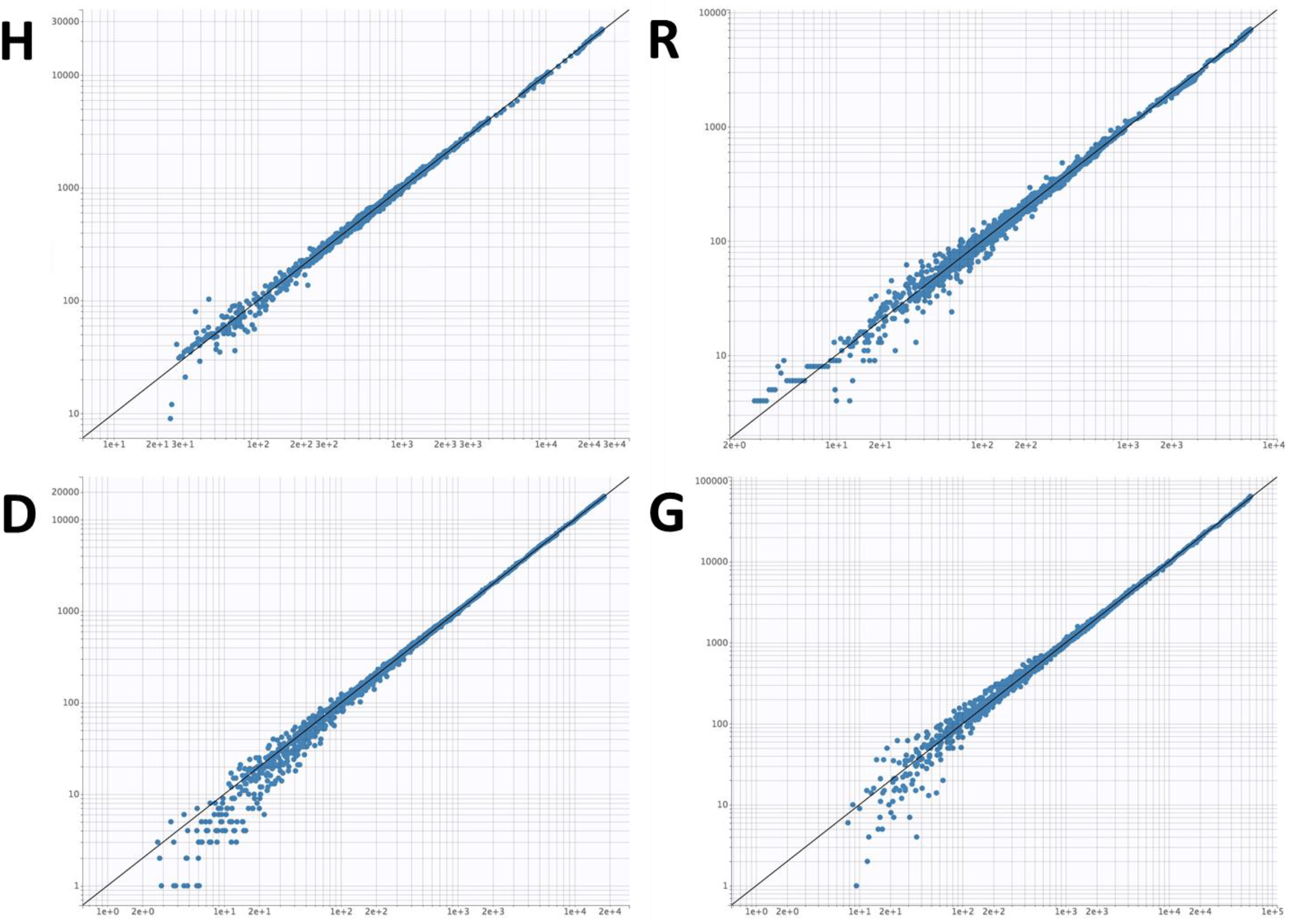
Model-fitted vs. observed values of hospitalizations in non-ICU services (H), in ICU (R), deaths in hospital (D) and returns at home (G) in log-scale.

**Figure 4.**
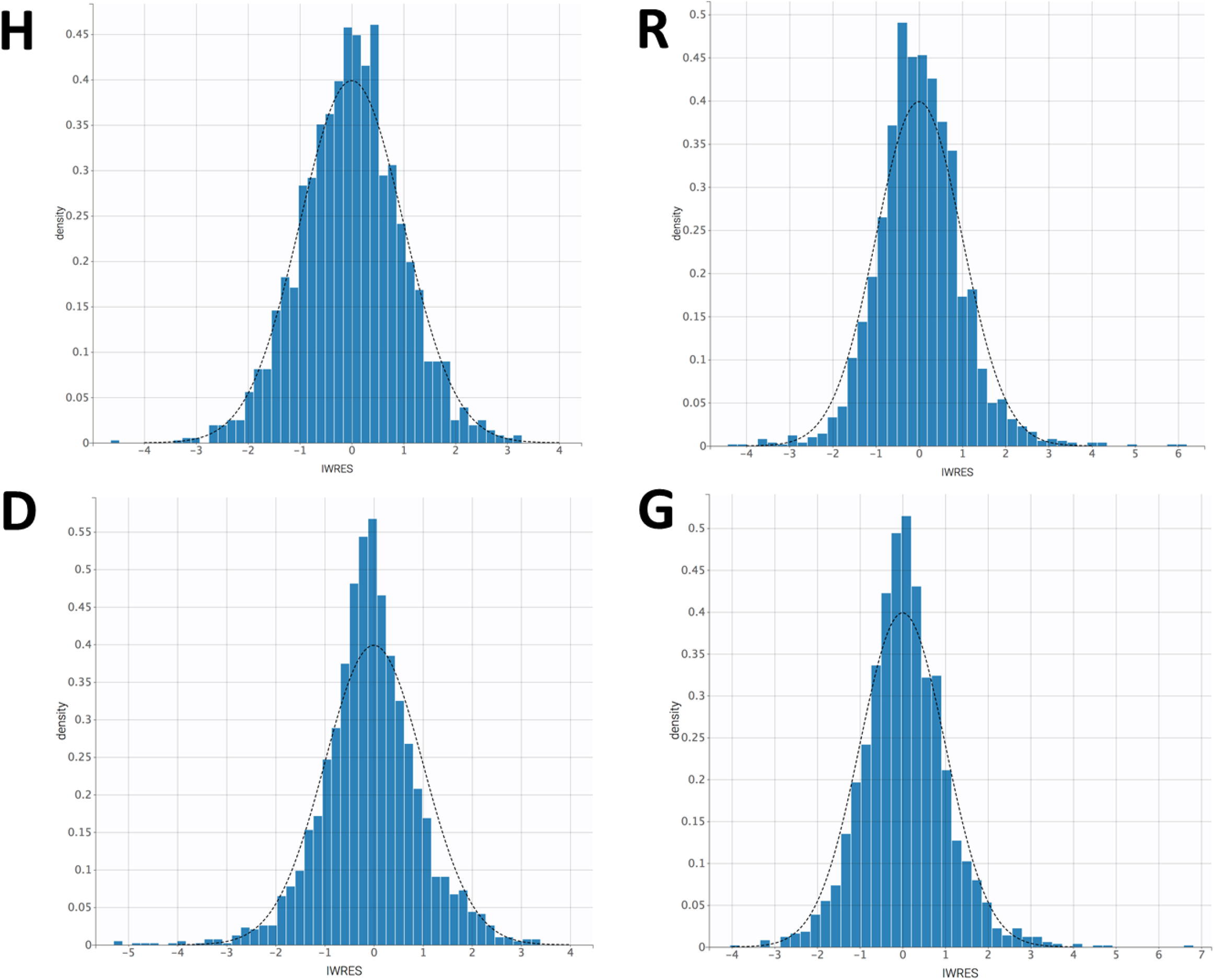
Weighted “individual” residuals vs. Gaussian law.

If the inter-region variability of parameters related to disease itself was low (< 40% for t_first_, N_0_ and k_I_), this variability was much higher for parameters related to hospital flows (between 40% and 95%), and surprisingly high for lockdown effect (130% for α). The lower the α, the higher the lockdown effect on k_I_ (table 2). For instance, lockdown was very effective in Ile-de-France (α=0.013), while less efficient in Grand-Est (α=0.52) or Provence-Alpes-Côte d’Azur (α=0.38). On the global France data, a poor effect of lockdown was observed (α=0.89). Of note, no inter-region correlation was found between parameters related to disease and related to lockdown (α, t_effect_).

**Table 2.** Parameter estimates in whole France, and all regions and some sub-regions.

| Region | $t_{\text{first}}$ | $N_0$ | $N_0^*$ | $k_I$ | $k_{HD}$ | $k_{DH}$ | $k_H$ | $k_{H2}$ | $k_{G1}$ | $k_R$ | $k_{RD}$ | $k_{G2}$ | $k_{DR}$ | $\alpha$ | Lockdown<br>effect delay | Population |
| --- | --- | --- | --- | --- | --- | --- | --- | --- | --- | --- | --- | --- | --- | --- | --- | --- |
| France | 40 | 108658 | 162 | 0.33 | 0.0080 | 0.00035 | 0.028 | 0.018 | 0.087 | 0.26 | 0.33 | 0.071 | 0.18 | 0.15 | 17 | 67064000 |
| IDF | 47 | 40798 | 332 | 0.36 | 0.0095 | 0.000028 | 0.035 | 0.019 | 0.074 | 0.33 | 0.59 | 0.054 | 0.15 | 0.16 | 15 | 12278200 |
| Paris | 49 | 9357 | 428 | 0.35 | 0.0076 | 0.0013 | 0.050 | 0.028 | 0.057 | 0.40 | 0.62 | 0.042 | 0.14 | 0.31 | 13 | 2187530 |
| H-D-Seine | 54 | 6333 | 395 | 0.36 | 0.0113 | 0.0001 | 0.036 | 0.019 | 0.080 | 0.45 | 0.72 | 0.052 | 0.14 | 0.33 | 14 | 1603270 |
| V-D-Marne | 52 | 5969 | 430 | 0.34 | 0.0132 | 0.0016 | 0.052 | 0.025 | 0.056 | 0.27 | 0.51 | 0.044 | 0.22 | 0.80 | 15 | 1387930 |
| S-S-Denis | 53 | 5257 | 324 | 0.33 | 0.0100 | 0.000031 | 0.040 | 0.018 | 0.074 | 0.74 | 2.06 | 0.036 | 0.25 | 0.56 | 19 | 1623110 |
| Essonne | 53 | 3495 | 270 | 0.31 | 0.0075 | 0.000014 | 0.038 | 0.020 | 0.041 | 0.37 | 0.42 | 0.068 | 0.15 | 0.46 | 12 | 1296130 |
| Yvelines | 53 | 3454 | 240 | 0.33 | 0.0125 | 0.00019 | 0.039 | 0.020 | 0.083 | 0.85 | 1.24 | 0.061 | 0.19 | 0.15 | 14 | 1438270 |
| Val-Oise | 54 | 3951 | 322 | 0.37 | 0.0197 | 0.0012 | 0.049 | 0.029 | 0.135 | 0.32 | 0.51 | 0.079 | 0.25 | 0.59 | 4 | 1228620 |
| Seine-Marne | 58 | 3631 | 259 | 0.35 | 0.0044 | 0.00017 | 0.028 | 0.015 | 0.111 | 0.98 | 1.58 | 0.094 | 0.14 | 0.51 | 15 | 1404000 |
| Grand-Est | 45 | 16773 | 304 | 0.34 | 0.0091 | 0.0045 | 0.033 | 0.021 | 0.072 | 0.42 | 0.51 | 0.069 | 0.40 | 0.40 | 15 | 5511750 |
| Bas-Rhin | 54 | 3478 | 309 | 0.41 | 0.0086 | 0.0043 | 0.024 | 0.021 | 0.065 | 0.33 | 0.22 | 0.076 | 0.29 | 0.36 | 14 | 1125560 |
| Haut-Rhin | 45 | 4630 | 606 | 0.32 | 0.0114 | 0.00084 | 0.038 | 0.020 | 0.092 | 0.50 | 1.20 | 0.041 | 0.81 | 0.11 | 6 | 764030 |
| Moselle | 54 | 3790 | 363 | 0.37 | 0.0047 | 0.0050 | 0.036 | 0.018 | 0.056 | 0.24 | 0.58 | 0.051 | 0.29 | 0.15 | 10 | 1043520 |
| GE-autres | 51 | 6474 | 251 | 0.34 | 0.0117 | 0.0024 | 0.051 | 0.029 | 0.082 | 0.78 | 1.27 | 0.083 | 0.20 | 0.58 | 9 | 2578640 |
| H-D-France | 37 | 13191 | 221 | 0.24 | 0.0081 | 0.00076 | 0.028 | 0.022 | 0.075 | 0.95 | 0.93 | 0.095 | 0.23 | 0.17 | 14 | 5962660 |
| Nord | 51 | 6516 | 250 | 0.29 | 0.0148 | 0.0024 | 0.030 | 0.035 | 0.075 | 0.97 | 0.61 | 0.089 | 0.21 | 0.18 | 12 | 2604360 |
| HDF-autres | 21 | 15025 | 447 | 0.21 | 0.0102 | 0.37 | 0.055 | 0.038 | 0.035 | 1.45 | 0.54 | 0.151 | 0.17 | 0.39 | 17 | 3358300 |
| Auvergne-RA | 48 | 17675 | 220 | 0.33 | 0.0142 | 0.0061 | 0.039 | 0.027 | 0.086 | 0.25 | 0.15 | 0.086 | 0.26 | 0.18 | 8 | 8032380 |
| Rhone | 51 | 4867 | 1064 | 0.33 | 0.0115 | 0.0052 | 0.035 | 0.024 | 0.061 | 0.25 | 0.16 | 0.075 | 0.28 | 0.23 | 8 | 457392 |
| ARA-autres | 48 | 20577 | 272 | 0.31 | 0.0156 | 0.0064 | 0.046 | 0.032 | 0.098 | 0.32 | 0.20 | 0.094 | 0.20 | 0.17 | 9 | 7574990 |
| PACA | 33 | 21868 | 433 | 0.28 | 0.0068 | 0.00015 | 0.065 | 0.068 | 0.043 | 0.33 | 0.15 | 0.450 | 0.07 | 0.42 | 12 | 5055650 |
| B-D-Rhone | 46 | 6124 | 303 | 0.29 | 0.0106 | 0.00011 | 0.054 | 0.025 | 0.097 | 0.25 | 0.26 | 0.153 | 0.07 | 0.36 | 10 | 2024160 |
| PACA-autres | 35 | 6854 | 226 | 0.27 | 0.0054 | 0.00010 | 0.069 | 0.077 | 0.039 | 2.43 | 1.01 | 0.409 | 0.09 | 0.51 | 15 | 3031490 |
| Bourgogne-FC | 46 | 6011 | 216 | 0.32 | 0.0178 | 0.0035 | 0.058 | 0.035 | 0.115 | 0.49 | 0.70 | 0.071 | 0.23 | 0.69 | 5 | 2783040 |
| Centre-VDL | 57 | 4261 | 167 | 0.33 | 0.0168 | 0.0043 | 0.031 | 0.033 | 0.054 | 0.82 | 0.44 | 0.120 | 0.26 | 0.29 | 12 | 2559070 |
| Occitanie | 48 | 7249 | 122 | 0.29 | 0.0083 | 0.020 | 0.043 | 0.030 | 0.082 | 1.02 | 0.31 | 0.086 | 0.23 | 0.14 | 9 | 5924860 |
| Aquitaine | 55 | 8807 | 147 | 0.35 | 0.0148 | 0.0079 | 0.044 | 0.040 | 0.074 | 0.33 | 0.12 | 0.090 | 0.29 | 0.17 | 7 | 5999980 |
| Pays-Loire | 51 | 10926 | 287 | 0.30 | 0.0087 | 0.00069 | 0.042 | 0.042 | 0.045 | 0.27 | 0.17 | 0.233 | 0.13 | 0.19 | 9 | 3801800 |
| Normandie | 57 | 6236 | 189 | 0.34 | 0.0128 | 0.00025 | 0.029 | 0.034 | 0.074 | 0.20 | 0.15 | 0.114 | 0.18 | 0.09 | 9 | 3303500 |
| Bretagne | 36 | 7358 | 220 | 0.23 | 0.0109 | 0.00055 | 0.057 | 0.046 | 0.059 | 0.46 | 0.29 | 0.116 | 0.23 | 0.27 | 14 | 3340380 |
| Corse | 49 | 367 | 107 | 0.28 | 0.0080 | 0.00065 | 0.046 | 0.020 | 0.088 | 0.27 | 0.51 | 0.064 | 0.13 | 0.22 | 7 | 344679 |

**Legends:** $N_0^*$ : number of patients bound to be hospitalized per 100,000 capita. Date of first admission is an estimate of the date where the first patient was hospitalized such as $H_1(t = t_{\text{first}}) = 1$ , lockdown after delay is the number of days after which lockdown effect is detected since lockdown date on March 17<sup>th</sup> of 2020. Other abbreviations are found in the text.

The time for which the first patient was hospitalized in France corresponds to the initiation of the system effect such as H_1_(t=t_first_) = 1, at day 47, which corresponds to the February 17^th^ of 2020. French lockdown started one month later, on March 17^th^ of 2020 (day 76); the effect of lockdown was detectable 12 days after this date in whole France (day 88, table 2), variable among (sub)regions.

Simulations suggested that lockdown did not substantially decrease either hospital occupancy or deaths in hospital. They also suggested that lockdown effect would have been increased if started before its actual date. Indeed, hospital occupancy and deaths in hospital would have been both decreased by 30% and 85% in average if lockdown was started 20 and 30 days before this date, respectively. (figure 5).

**Figure 5.**
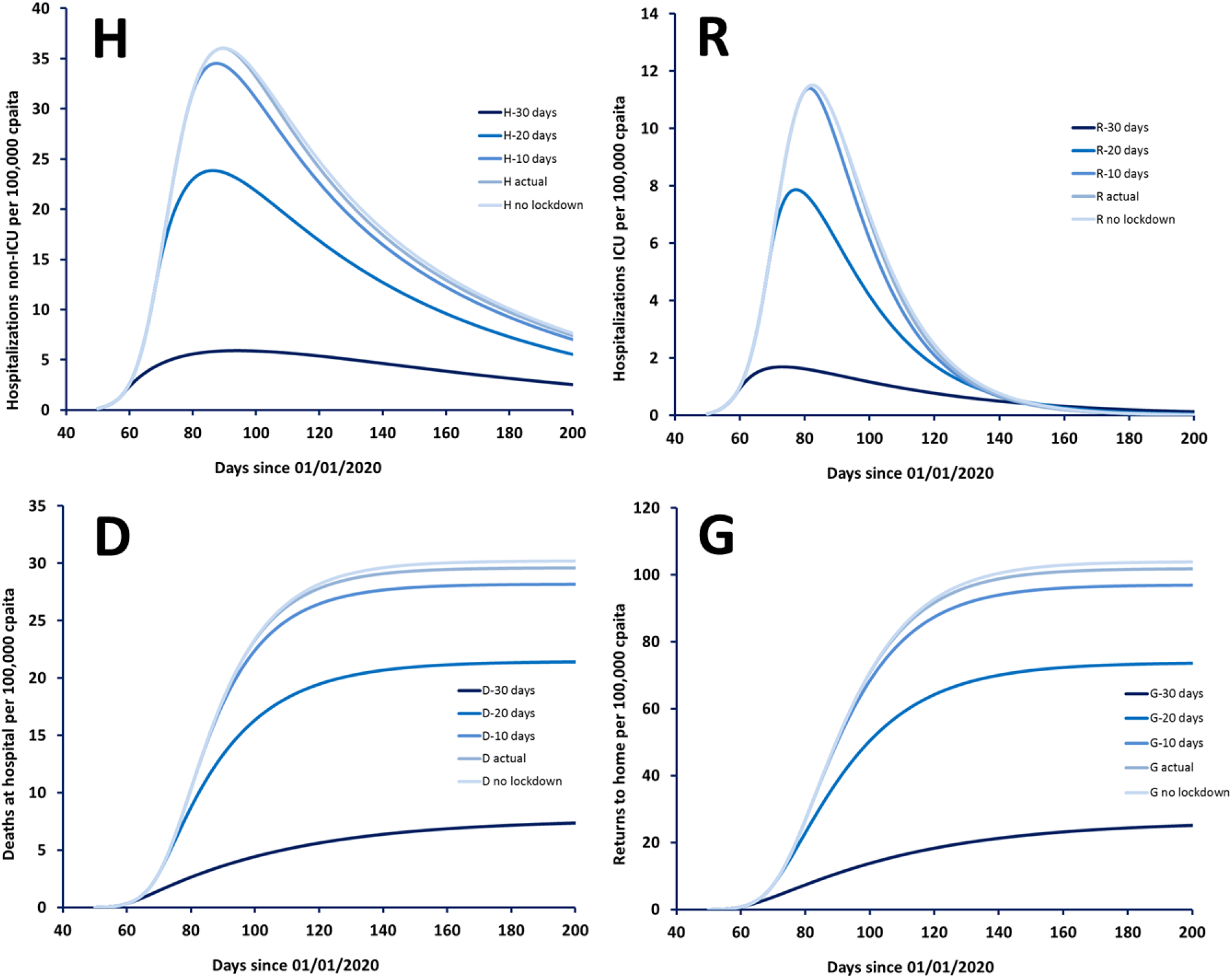
Simulations of hospitalizations in non-ICU services (H), in ICU (R), deaths in hospital (D) and returns at home (G) for different lockdown strategies. The value of N_0_ was converted as its value per 100,000 capita (N_0*_=170). Lockdown effect was detectable at day 88, i.e. 12 days after start of lockdown.

## 4 Discussion

This work has focused on available data to estimate the actual effect of lockdown on hospitalization burden in regions and sub-regions of France. This model allowed an accurate description of patient flows between non-ICU services, ICU, deaths at hospital and return to home. Adding the effect of lockdown allowed an even better description of hospital data.

A previous work suggested that French lockdown would have prevented almost 600,000 hospital admissions, including almost 150,000 in ICU [9]. However, this work has been criticized, because of a possible over-estimation of hospital burden [12]. The principle of lockdown itself remains controversial. Introduced for the first time in human history in 2020 instead of isolation or quarantine, it may lead to a large social cost while being less efficient than isolation or quarantine measures [13]. To deal with this controversy, the present hospital flow model was attempted to either validate or invalidate the following hypothesis: “lockdown has decreased hospital burden in France”. This necessitated a sound description of patient input/flows/output in hospital.

This study showed a significant effect of lockdown on hospitalization rate (k_I_), with a global decrease of k_I_ by thrice. However, the consequences of lockdown appeared to be minor: simulations made from the hospital flow model suggested that lockdown have had no significant influence on hospital burden and would have saved less than 1 life per 100,000 capita, compared to the 22 per 100,000 who have died until the day May 22^nd^ [14]. There may be at least three explanations: first, the effect of lockdown is delayed in time (detected 12 days after start of lockdown in average), second, hospital occupancy was already high at this date and, last, but not least, the risk of infection may have already started to decline [14]. Nevertheless, the effect of lockdown, as well as patient flow parameters, was found to be very variable among regions and sub-regions of France. This variability may be due, at least in part, to population density in each region, different hospital cares, different population characteristics (inter-region differences in mean age, co-morbidities, etc.), and/or to the respect of mitigation strategies.

If an intrinsic effect of lockdown measures seems indisputable, one could question the date of establishment. Indeed, simulations showed that both hospital occupancy and deaths would not have been significantly different if no lockdown was established, but they might have been dramatically decreased if lockdown had been started 20 or 30 days started before the official date of application. Nevertheless, at these days, the first death from COVID-19, and the first case were respectively recorded.

This work has nevertheless limitations. First, all French sub-regions were not considered, in order to keep N_0_ sufficiently high in each region unit and to avoid very long time of computation. In particular, overseas territories were not considered, except for Corse. Second, even if accurate values of model parameters were obtained in each region, notably the time of the first hospitalized patient, no spatial description of disease spread was made. Third, this model does not pretend to be an epidemic model: the hospital admission growth was described using a Kermack-McKendrick input function, which may appear oversimplistic. This also concerns the “on/off” model for lockdown effect, which may not account for the complexity of the real effect of lockdown. Therefore, this model cannot test lockdown ending strategies and their consequences on the risk and amplitude of epidemic rebounds. To describe or forecast these rebounds, a mechanistic epidemic model as Roux et al. [9] could elegantly replace the present input function. Last but not least, this model is “macroscopic”, i.e. does not account for patient specificities (as age distribution or proportion of comorbidities). Taking into account these factors may improve the description of inter-region variability [14]. Given these limitations, the lockdown effect should be considered with caution, this study providing trends more than a sound quantification.

In conclusion, the present model has shown an intrinsic effect of lockdown to decrease hospital burden. However, the efficiency of lockdown may have been increased if established sooner. This may raise important questions, notably regarding social acceptability of these rigorous measures that would have imposed while no strong signal of danger was present.

## Data Availability

Data are available on official sources

## Notes

### Competing Interest Statement

The authors have declared no competing interest.

### Funding Statement

No funding for this study

### Author Declarations

These data are public data, obtained in official sites.

## References

1. COVID-19 Coronavirus Pandemic. 2020. https://www.worldometers.info/coronavirus/

2. Neil M Ferguson et al. Report 9: Impact of non-pharmaceutical interventions (NPIs) to reduce COVID-19 mortality and healthcare demand, 2020. https://www.imperial.ac.uk/media/imperial-college/medicine/mrc-gida/2020-03-16-COVID19-Report-9.pdf

3. COVID-19 Pandemic Lockdowns (Wikipedia). https://en.wikipedia.org/wiki/COVID-19_pandemic_lockdowns.

4. Suivi de l’épidémie de COVID-19 en France. 2020. https://dashboard.covid19.data.gouv.fr/

5. Patrick GT Walker et al. The Global Impact of COVID-19 and Strategies for Mitigation and Suppression, 2020 https://www.imperial.ac.uk/media/imperial-college/medicine/sph/ide/gida-fellowships/Imperial-College-COVID19-Global-Impact-26-03-2020v2.pdf

6. Fizza Farooq et al. Effect of Lockdown on the spread of COVID-19 in Pakistan. 2020. https://arxiv.org/ftp/arxiv/papers/2005/2005.09422.pdf

7. Osmar Pinot Neto et al. Mathematical model of COVID-19 intervention in Sao Paulo – Brazil. 2020. https://arxiv.org/ftp/arxiv/papers/2005/2005.09426.pdf.

8. R. Adhikari et al. Inference, prediction and optimization of non-pharmaceutical interventions using compartment models: the PyRoss library. 2020. https://arxiv.org/pdf/2005.09625.pdf

9. Jonathan Roux et al. COVID-19: One-month impact of the French lockdown on the epidemic burden. 2020. https://www.ehesp.fr/wp-content/uploads/2020/04/Impact-Confinement-EHESP-20200322v1-1.pdf

10. Diane R Mould, et al. Basic concepts in population modeling, simulation, and model-based drug development. CPT Pharmacometrics Syst Pharmacol. 2012;1:e6

11. Diane R Mould et al. Basic Concepts in Population Modeling, Simulation, and Model-Based Drug Development-Part 2: Introduction to Pharmacokinetic Modeling Methods CPT Pharmacometrics Syst Pharmacol. 2013;2:e38

12. Tribune : Etude critique d’une modélisation des effet du confinement. Journal international de médecine, 05/02/2020 https://www.jim.fr/medecin/jimplus/e-docs/etude_critique_dune_modelisation_des_effets_du_confinement182808/document_edito.phtml

13. M. Zelmat. Facing COVID-19 by the confinement :EBM, “MBM” or “SBM” ? 2020. https://papers.ssrn.com/sol3/papers.cfm?abstract_id=3600511

14. Juanjuan Zhang. Fundamental principles of epidemic spread highlight the immediate need for large-scale serological surveys to assess the stage of the SARS-CoV-2 epidemic. 2020. https://www.reddit.com/r/COVID19/comments/fonnt8/fundamental_principles_of_epidemic_spread/

